# Echocardiography-Based, Artificial Intelligence-Enabled Electrocardiography (AI-ECG) for Diastolic Hemodynamics Phenotyping in Acute Heart Failure (AHF)

**DOI:** 10.64898/2026.03.05.26347763

**Authors:** Yee Weng Wong, Muhannad A. Abbasi, Eunjung Lee, Gal Tsaban, Zachi Attia, Paul A. Friedman, Peter A. Noseworthy, Francisco Lopez-Jimenez, Horng H. Chen, Grace Lin, Luis R. Scott, Omar F. Abou Ezzeddine, Jae K. Oh

## Abstract

**Background:** Acute heart failure (AHF) exhibits marked heterogeneity in diastolic hemodynamics, yet comprehensive echocardiographic assessment of diastolic function (DF) and filling pressure (FP) is often infeasible. We evaluated whether artificial intelligence–enabled electrocardiography (AI-ECG) could provide scalable DF grading and FP estimation in hospitalized AHF patients.

**Methods:** We retrospectively studied adults hospitalized for AHF across Mayo Clinic sites (2013–2023) who received ≥1 dose of intravenous loop diuretic and had paired 12-lead ECG and TTE. The previously validated AI-ECG DF model was applied without retraining to generate four DF grades and a continuous FP probability. Clinical outcomes were all-cause mortality and heart failure rehospitalization. Associations with clinical severity markers and echocardiographic indices were examined. Kaplan–Meier survival analysis and adjusted multivariable Cox proportional hazards models were performed. Exploratory analyses examine the kinetics of change in FP probability and impact on mortality.

**Results:** Among 11,513 patients (median age 75 years, 39% female), AI-ECG DF grading was feasible in 100%, whereas echocardiographic DF was indeterminate in 44% of clinically eligible patients. In 2,582 patients with determinate echocardiographic DF, AI-ECG FP probability discriminated TTE Grade 2–3 dysfunction with AUC 0.85 (95% CI 0.83 – 0.86). Higher AI-ECG DF grades were associated with higher comorbidity burden, worse NYHA class, elevated NT-proBNP, higher MAGGIC scores, elevated PCWP, and more advanced structural remodeling. After multivariable adjustment, AI-ECG DF remained independently associated with mortality (hazard ratio [HR] 1.25, 95% CI 1.16–1.35 for Grade 2; HR 1.44, 95% CI 1.33–1.56 for Grade 3 versus Normal/Grade 1). Combining AI-ECG DF with MAGGIC scores yielded ordered risk gradients, with highest mortality in patients with both high MAGGIC and Grade 2–3 DF. Among patients with serial ECGs, improvement in FP probability was independently associated with lower mortality (HR 0.85, 95% CI 0.79–0.91), whereas worsening did not show a consistent adverse gradient beyond baseline DF.

**Conclusions:** In a large, geographically diverse AHF cohort, AI-ECG DF grading was universally feasible, correlated with established hemodynamic severity markers, and provided independent prognostic information beyond established risk factors, supporting its role as a pragmatic, scalable diastolic biomarker in AHF.

**CLINICAL PERSPECTIVE:** *What Is New?:*

- In 11,513 hospitalized acute heart failure (HF) patients, artificial intelligence-enabled electrocardiography provided diastolic function grading in 100% of patients from a single 12-lead ECG without requiring additional clinical variables, compared with 56% feasibility for guideline-based echocardiography grading.
- AI-ECG diastolic function grades correlated with established marker of severity (NYHA functional class, NT-proBNP, MAGGIC risk scores, and pulmonary capillary wedge pressure) and remained independently associated with both mortality and HF rehospitalization after multivariable adjustment.
- Serial AI-ECG measurements identified post-discharge filling pressure trajectories, with improvement independently associated with 15% lower mortality, a first demonstration that longitudinal ECG assessment can track post-discharge hemodynamic recovery.

*What Are the Clinical Implications?:*

- AI-ECG transforms the universally obtained 12-lead ECG into an actionable hemodynamic biomarker that addresses the critical gap when echocardiographic diastolic function assessment is indeterminate or unavailable in acute HF patients.
- Despite markedly different hemodynamic severity and long-term outcomes across AI-ECG diastolic function grades, hospitalization length of stay did not differ, suggesting advanced diastolic dysfunction represents occult risk not easily recognized during routine acute care and highlighting the need for improved post-discharge risk stratification.
- The continuous filling pressure probability metric enables longitudinal monitoring of post-discharge hemodynamic status using serial routine ECGs, potentially identifying patients requiring intensified follow-up or specialist referral.

## Background

Acute heart failure (AHF) hospitalization is associated with increased post-discharge adverse events,^1^ highlighting the importance of individualized risk assessment to guide decongestion and follow-up. Identifying advanced diastolic dysfunction (DF) and increased left ventricular (LV) filling pressures (FP) is important, as these changes lead to congestion and adverse outcomes in all ejection fractions (EF)^2, 3^. Additionally, a restrictive filling profile is independently associated with increased mortality, highlighting the prognostic value of DF beyond systolic function.^4^

Echocardiography is the reference non-invasive modality for DF assessment and LV FP estimation.^5^ Application of guideline-based DF grading at AHF discharge independently stratifies post-discharge risks, highlighting the clinical relevance of diastolic hemodynamics in the acute-care continuum.^6^ However, universal echocardiography is operationally infeasible, and DF adjudication is frequently limited. In large multicenter cohorts, guideline DF grading is indeterminate in at least one-fifth of studies^7^, and common conditions (tachycardia, atrial fibrillation, suboptimal windows) limit algorithm application.^5^

Symptom-based triage strategies anchored to dyspnea are inherently limited in AHF. Prior observation demonstrated that many patients hospitalized with AHF report fatigue, peripheral edema, or ascites as their predominant presenting symptom rather than dyspnea, underscoring the heterogeneity of clinical presentation.^8^ Reliance on dyspnea alone therefore risks both under- and over-diagnosis of AHF.^9^ These observations highlight the critical need for objective, scalable, and noninvasive methods for congestion assessment that transcend symptom-based frameworks and allow timely identification of elevated FP.

Artificial-intelligence enabled electrocardiography (AI-ECG) offers a pragmatic, deployable supplemental biomarker.^10^ Using a 12-lead ECG, a deep-learning model can grade DF and estimate elevated FP probability with high discrimination. The model outputs are independently associated with mortality and demonstrate performance comparable to echocardiography, including in patients with indeterminate echocardiographic DF.^11^ Because ECG acquisition is ubiquitous, rapid, and inexpensive, AI-ECG can extend access to noninvasive hemodynamic risk stratification in AHF, potentially addressing therapeutic inertia in real-world acute-care workflows.

Building on these observations, this retrospective multisite study evaluated the utility of AI-ECG for noninvasive hemodynamic phenotyping in AHF. The objectives were to: (i) assess the feasibility of AI-ECG–derived DF grading in a geographically diverse AHF population; (ii) examine the association between AI-ECG DF Grades and established measures of disease severity; (iii) evaluate the prognostic value of AI-ECG DF Grades beyond conventional risk factors; and (iv) in exploratory analyses, to assess the prognostic implications of changes in AI-ECG–predicted FP probability during post-discharge follow-up.

## METHODS

### Study Design and Population

We conducted a retrospective cohort study of adults (≥18 years) hospitalized for AHF at Mayo Clinic hospitals in Minnesota, Arizona, and Florida, including Mayo Clinic Health System sites. Eligible patients had hospitalization ≥24 hours with clinically documented AHF (confirmed by ICD coding), received at least one dose of intravenous (IV) loop diuretic therapy, and had both 12-lead ECG and TTE performed during hospitalization. Patients were excluded if ECG or TTE data were unavailable or incomplete, or if they had been included in the derivation or validation cohorts used to train the AI-ECG model. The study was approved by the Mayo Clinic Institutional Review Board with waiver of informed consent.

### AI-ECG DF Model Architecture and Output Interpretation

This study applied the previously developed AI-ECG diastolic function (DF) model by Lee et al.^11^ without retraining, to an independent clinical cohort.

The original model was trained on over 200,000 paired ECG and TTE studies, with echocardiography-based DF grades (Normal, Grade 1–3) derived using the Mayo Clinic 2020 algorithm^12^, a slight simplification of the 2016 ASE/EACVI guidelines, with close alignment with recent 2025 update (Supplemental Figure 1).^5,11^ Ten-second, 12-lead ECGs sampled at 500 Hz were structured as time × lead matrices and divided into five non-overlapping 2-second segments. The model architecture was a modified ResNet-18 convolutional neural network (a deep learning pattern-recognition model originally designed for images, here adapted to ECG waveforms). In the original development study, the model demonstrated excellent discrimination for elevated FP (AUC 0.91) and Grade 3 diastolic dysfunction (AUC 0.94).

For interpretation, model outputs were reported both as four DF grades and as a continuous estimate of elevated FP. Each 10-second, 12-lead ECG generated four normalized probabilities summing to 1.0, corresponding to Normal, Grade 1, Grade 2, and Grade 3 DF. The DF grade assigned to an ECG was the class with the highest probability. For example, an ECG generating output of 0.10 for Normal, 0.10 for Grade 1, 0.30 for Grade 2, and 0.50 for Grade 3 would be assigned Grade 3 DF.

To construct a continuous FP measure, the probabilities for Grade 2 and Grade 3 were summed to yield an elevated-filling-pressure value (FP):

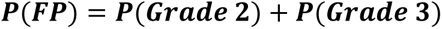

This continuous FP probability ranges from 0 to 1, where higher values reflect greater likelihood of elevated FP. For example, P(FP)=0.80 indicates high probability of elevated FP (typically Grade 3 DF), whereas P(FP)=0.20 suggests low likelihood (typically normal or Grade 1 DF). Of note, these outputs represent the model’s internal probability distribution and should not be interpreted as true clinical probabilities.

### Data Collection

We abstracted clinical and echocardiographic data from the electronic health record and Mayo Clinic Unified Data Platform. Baseline variables at hospitalization included demographic characteristics (age, sex, race), admission vital signs (systolic blood pressure, body mass index), medical history (prior HF, atrial fibrillation (AF), hypertension, diabetes mellitus (DM), coronary artery disease or previous myocardial infarction, chronic obstructive pulmonary disease, dementia and current tobacco use) and guideline-directed HF therapies prescriptions on admission, beta-blockers (BB), angiotensin-converting enzyme inhibitors (ACE-i) or angiotensin-receptor blockers (ARB), angiotensin receptor–neprilysin inhibitors (ARNI), mineralocorticoid-receptor antagonists (MRA), and sodium–glucose cotransporter-2 inhibitors (SGLT2-i).

Laboratory values at presentation included sodium, creatinine, hemoglobin, and N-terminal pro–B-type natriuretic peptide (NT-proBNP); NT-proBNP values were log-transformed for analysis. For invasive hemodynamic comparison, only pulmonary capillary wedge pressure (PCWP) obtained during invasive right-heart catheterization in the catheterization laboratory was included; bedside measurements were excluded.

Echocardiographic measurements were obtained from the transthoracic study performed during admission. Echocardiographic variables included LVEF, mitral E/e′ and E/A ratios, left atrial volume index (iLAV), and tricuspid regurgitation velocity. Using these, the patients’ diastolic function by echocardiography was categorized by the abovementioned algorithm. However, this was not available or indeterminate in a subset of patients.

Electrocardiographic measurements included cardiac rhythm, presence of left bundle-branch block (LBBB), PR, QRS, and QT intervals. When more than one ECG or echocardiogram was available, the test closest to the admission date was used. For follow-up ECG analyses, recordings obtained between day 7 and day 180 after discharge were selected.

### NYHA Functional Classification

Baseline NYHA functional class was independently adjudicated among consecutive patients with most complete echocardiographic characterization. A board-certified physician, blinded to AI-ECG and echocardiography-derived DF grades, reviewed admission documentation, and assigned NYHA class based on reported symptoms and functional capacity.

### MAGGIC Risk Score Calculation

The MAGGIC (Meta-Analysis Global Group in Chronic Heart Failure) risk score was calculated for the entire study cohort using baseline variables. The score is an established, integer-based mortality prediction model derived from more than 39,000 patients across 30 cohort studies and incorporates thirteen clinical variables (Supplemental Table 1).^13^ The MAGGIC scoring system accommodates missing variables within its model structure, allowing risk scores to be generated for all patients, including those without independently adjudicated NYHA classification. This feature permitted inclusion of the full cohort in mortality risk analyses.

### Clinical Outcomes

The primary outcome was all-cause mortality. Survival status and time-to-death were ascertained from the medical record and the National Death Index, with follow-up extended through the end of the study period. For patients discharged alive, survival time was measured from the index admission date to the date of death or last known follow-up. Heart-failure rehospitalization, defined by using the same clinical criteria applied to the index hospitalization (without the constraint of paired ECG-TTE requirement), was evaluated as a secondary outcome.

### AI-ECG DF Model Performance Evaluation

Computational repeatability was assessed by re-running the AI-ECG model on 100 randomly selected identical ECGs and comparing repeat FP probabilities. The model’s performance in detecting elevated LV FP was evaluated against TTE Grade 2–3 DF among patients with determinate DF grades. Discrimination of the continuous FP probability was evaluated using receiver operating characteristic (ROC) analysis with area under the curve (AUC). Sensitivity, specificity, PPV, and NPV were calculated using the previously published threshold of 0.264^11^, and a predefined specificity-optimized threshold of 0.6.

### Statistical Analysis

Analyses were prespecified and focused on the association between AI-ECG–derived DF grade and markers of disease severity, hemodynamics, and clinical outcomes.

Continuous variables were summarized as means ± standard deviations or medians with interquartile ranges; categorical variables as counts with percentages. Baseline characteristics were compared across AI-ECG DF categories (normal/Grade 1, Grade 2, Grade 3) using analysis of variance (ANOVA) or Kruskal–Wallis tests for continuous variables and χ² tests for categorical variables. Trend tests were applied for ordered variables.

MAGGIC risk score and resting pulmonary capillary wedge pressure were compared across DF categories using ANOVA with adjusted marginal means and pairwise contrasts. NT-proBNP distributions were compared using Kruskal–Wallis tests. NYHA class distributions were examined using contingency tables and χ² tests.

Time-to-event analyses for all-cause mortality used Kaplan–Meier estimates with log-rank tests. Cumulative incidence functions for HF rehospitalization and death within one year were estimated nonparametrically with Gray-type tests. Cox proportional hazards regression examined associations between DF grade and mortality. Multivariable models adjusted for age, sex, BMI, NYHA class, smoking status, history of MI, COPD, DM, hypertension, AF, hemoglobin, sodium, creatinine, systolic blood pressure, LVEF, baseline QRS duration, rhythm and LBBB status, prior HF, and baseline GDMT prescriptions. Covariates were selected a priori based on established associations with mortality in AHF and minimal missingness. Separate models replaced AI-ECG DF grade with echocardiographic DF grade. Survival analyses by combined risk categories that crossed MAGGIC strata (≥22 = high risk) with AI-ECG DF strata were conducted to characterize risk gradients.

Prespecified interaction analyses evaluated consistency of associations between AI-ECG DF and mortality across subgroups defined by echocardiographic DF determinacy, reduced ejection fraction (LVEF ≤40%), and sinus rhythm using interaction terms in multivariable Cox models. Additional models examined independent prognostic value of log-transformed NT-proBNP.

Longitudinal change in AI-ECG FP probability was assessed in patients with two qualifying ECGs: baseline and follow-up obtained 7–180 days post-discharge. Change over time was evaluated using repeated-measures ANOVA with time as within-subject factor and baseline DF grade as between-subject factor. Cox models incorporated change in FP probability (dichotomized as improved versus unchanged/worsened) alongside baseline DF grade and covariates from the primary model. The continuous change variable was then modeled using natural cubic splines (3 degrees of freedom) to characterize its functional association with mortality. Predictions were generated across the central distribution of the change variable (1st–99th percentile) with all covariates held at typical values, and HR were scaled to a reference of zero change. Nonlinearity was assessed by likelihood-ratio test comparing spline versus linear specifications. A dual-axis figure showed the spline-based adjusted hazard-ratio curve and the change variable’s distribution.

Proportional hazards assumptions were evaluated using Schoenfeld residuals. Functional form for continuous covariates was assessed using martingale residual plots. Significance threshold was P<0.05 for main effects; P<0.01 for interaction terms. Bonferroni or Holm adjustments were applied for post hoc comparisons. Analyses were conducted in R (R Foundation for Statistical Computing), selected figures were generated using GraphPad Prism.

## RESULTS

### Study Population

Among 220,236 adults with documented HF (2013–2023), 64,305 were hospitalized for AHF with research authorization and received at least one dose of intravenous loop diuretic. After excluding 43,958 patients without paired TTE and ECG, 20,347 patients formed the eligible cohort. TTE diastolic function (DF) was indeterminate in 8,931 patients (44%) and gradable in 11,416 patients (56%). To ensure independent validation, 8,834 patients whose data were previously used for AI-ECG DF model development were excluded from the TTE gradable group. The final analysis cohort comprised 11,513 patients: 8,931 with indeterminate TTE DF and 2,582 with determinate TTE DF. AI-ECG categorized patients as normal or Grade 1 diastolic function (30%), Grade 2 diastolic function (37%), and Grade 3 diastolic function (33%). The median time between baseline ECG and TTE was 0.8 days. The cohort was geographically diverse, representing nearly all U.S. states with substantial contributions from Midwest, South, and West regions (Figure 1).

**Figure 1.**
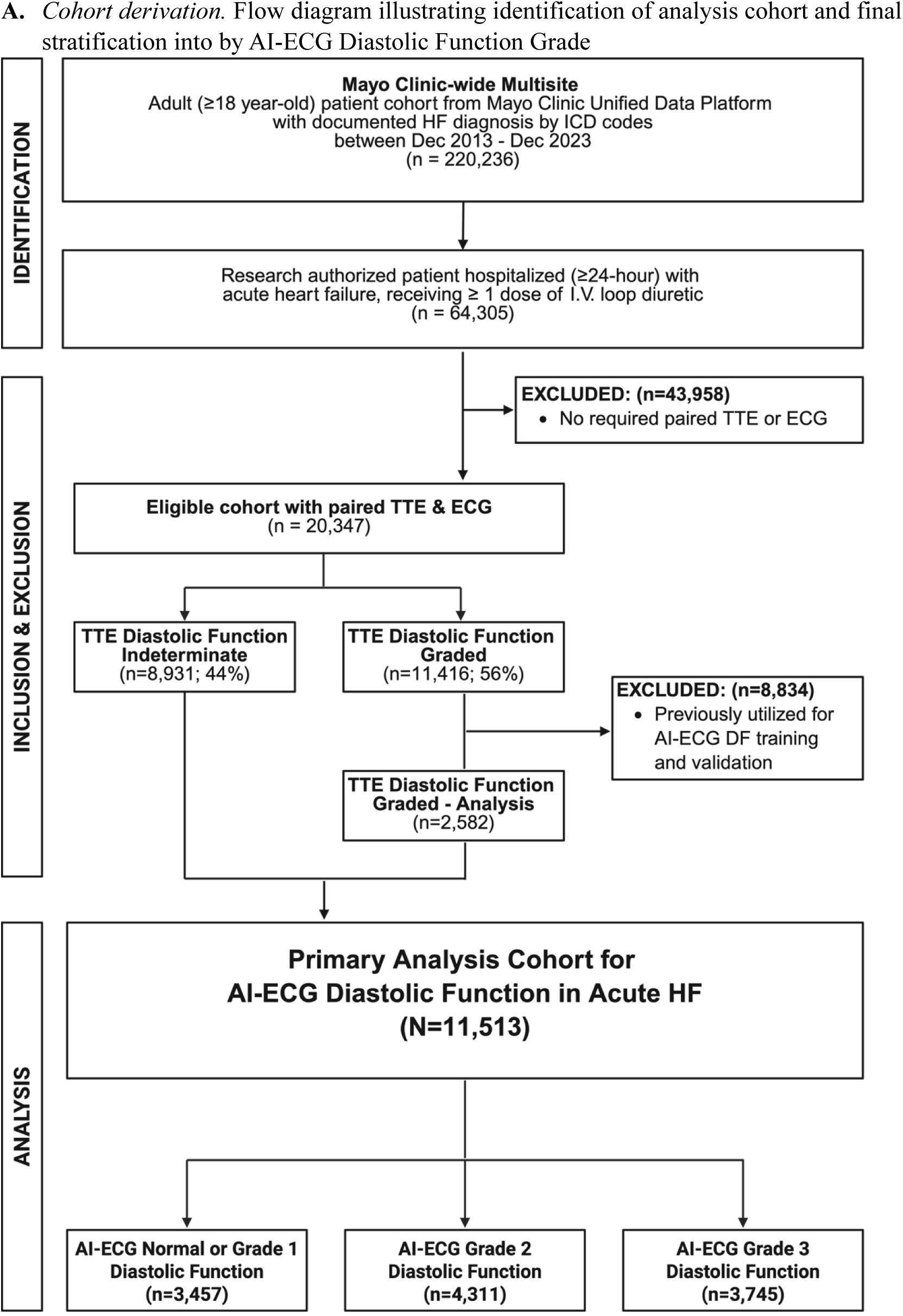
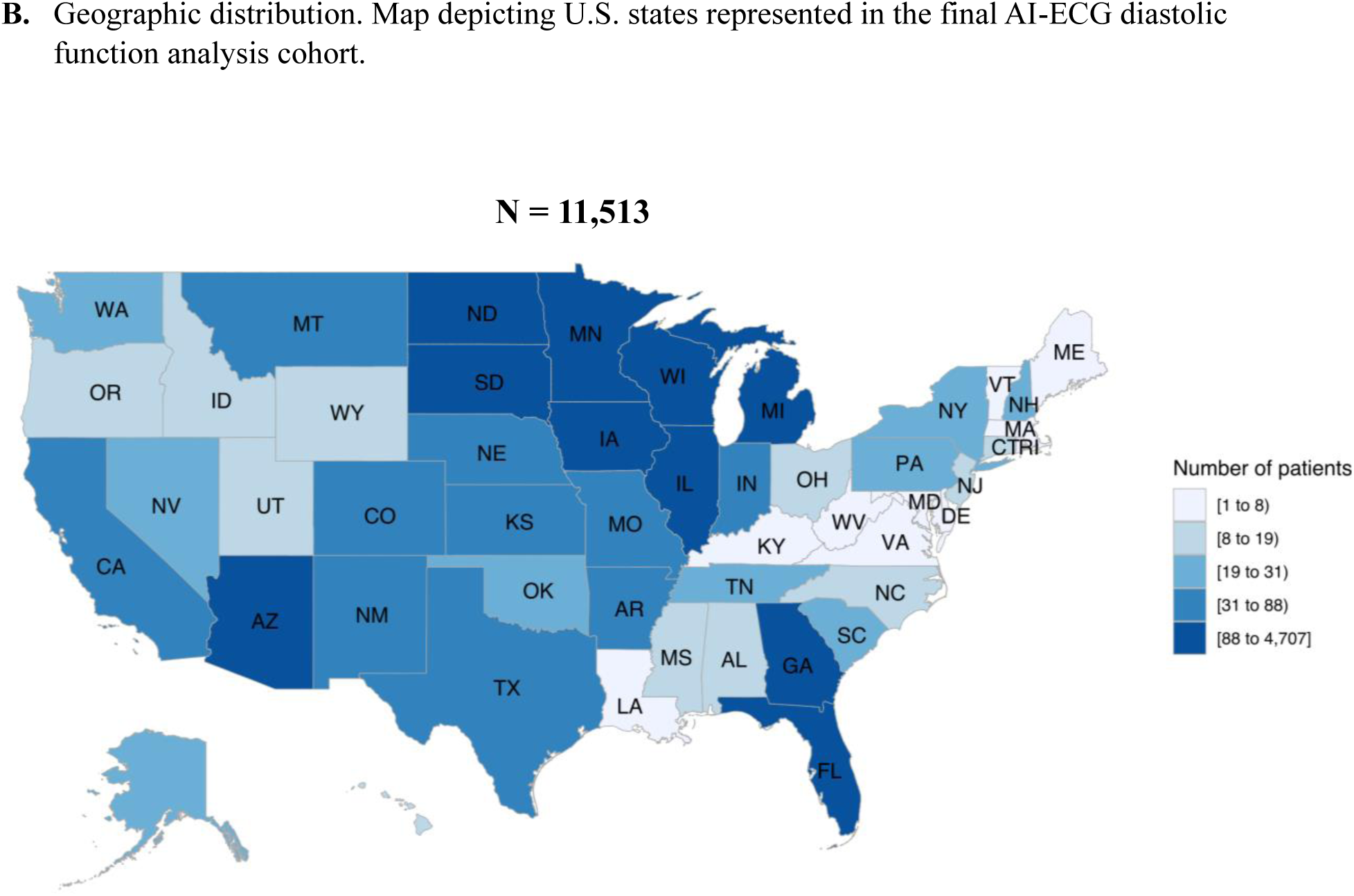
Study Cohort Assembly and Geographic Distribution of the AI-ECG Acute Heart Failure Population

### AI-ECG DF Model Performance Evaluation

Test–retest evaluation showed near-zero computational variability (differences in FP probability of 10⁻⁸ to 10⁻¹⁷), confirming deterministic, reproducible outputs. Among 2,582 patients with paired AI-ECG and echocardiography DF grades, AI-ECG FP probability discriminated TTE Grade 2–3 dysfunction with AUC 0.85 (95% CI 0.83–0.86). Using prespecified thresholds of 0.264 and 0.6 (prevalence 74.3%), sensitivity was 90.1% and 74.9%, specificity 57.7% and 79.7%, PPV 86.0% and 91.4%, and NPV 66.8% and 52.4%, respectively (Supplemental Figure 2). A confusion matrix for the complete cohort (including indeterminate TTE DF) annotated with observed death rates is provided (Supplemental Table 2).

### Baseline Characteristics

#### Demographic Characteristics

Patients with higher AI-ECG DF Grades were older, with mean age increasing from the normal/Grade 1 group to Grades 2 and 3. Women comprised 39.0% of the cohort, with the highest proportion in Grade 2. Most patients were White, with a higher proportion of Black patients in Grade 3 (Table 1).

**Table 1.**
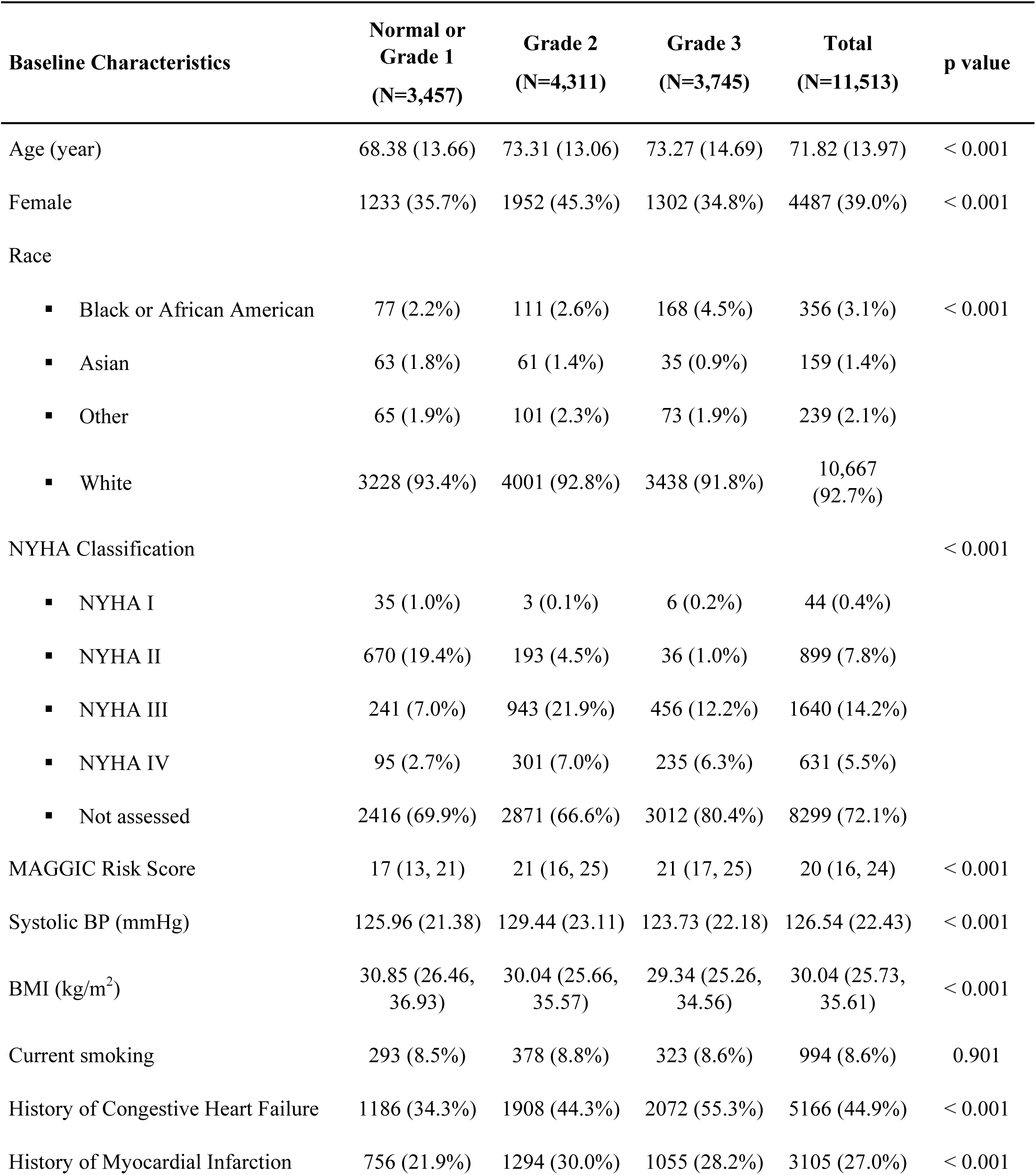
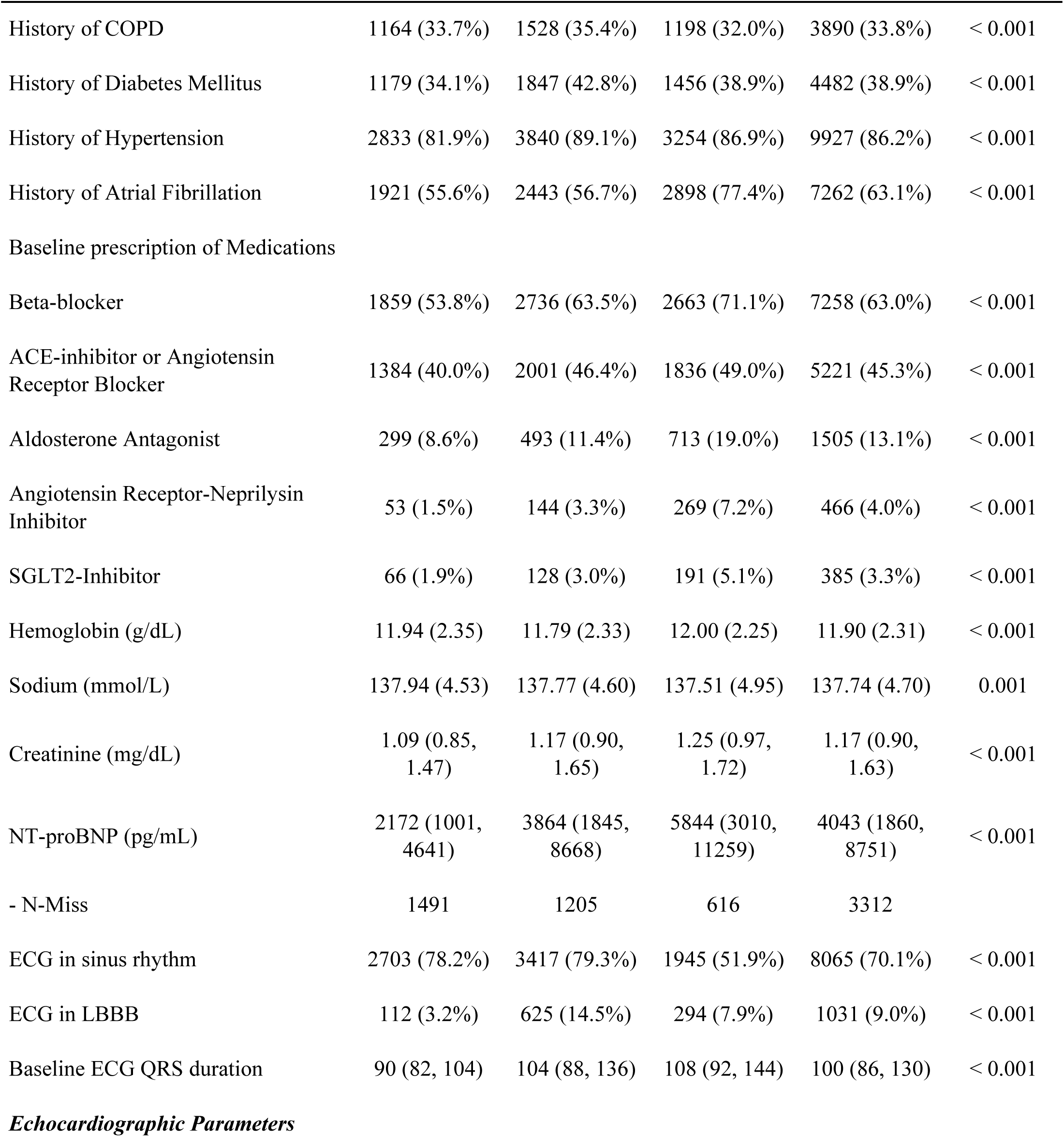
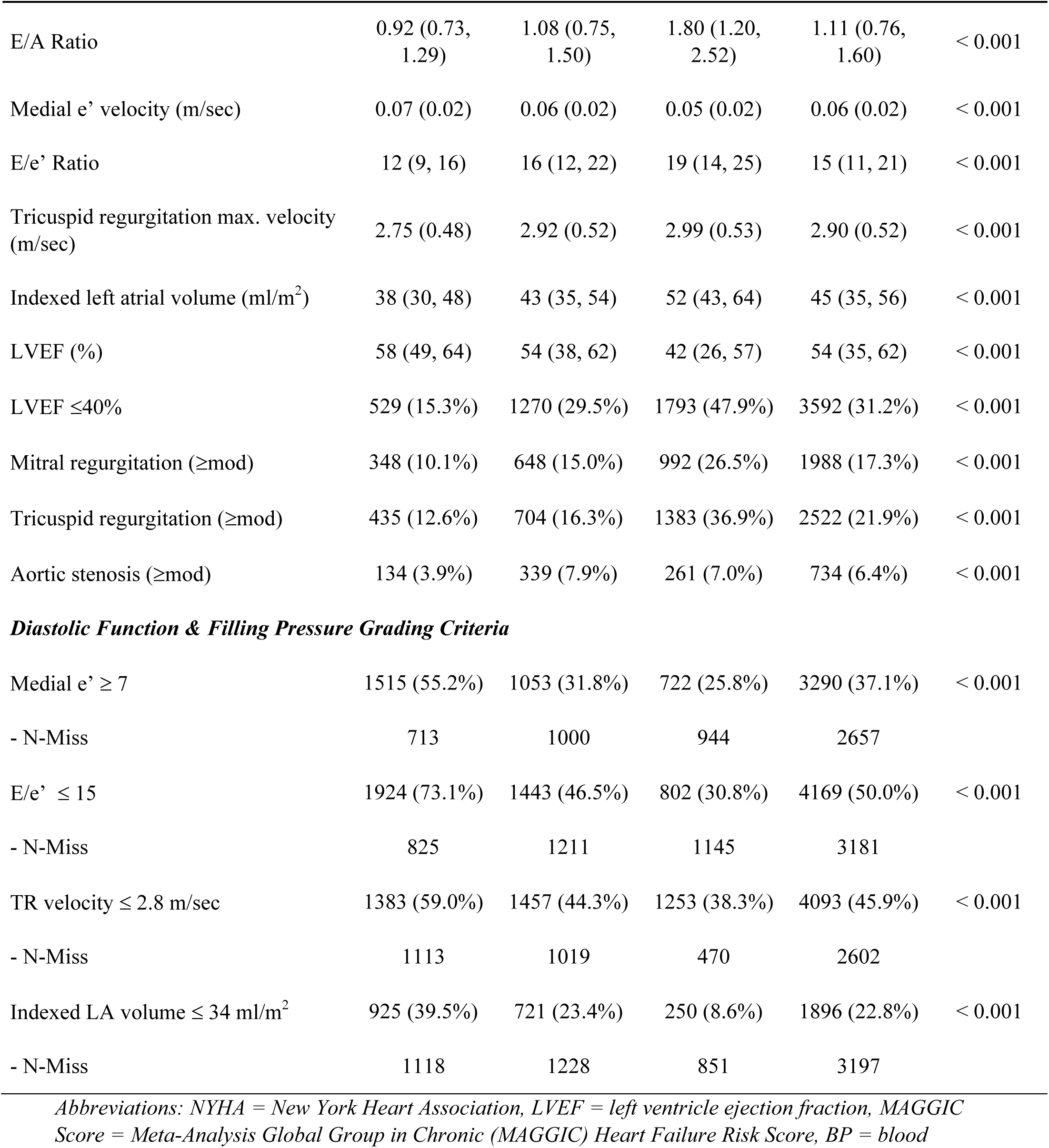
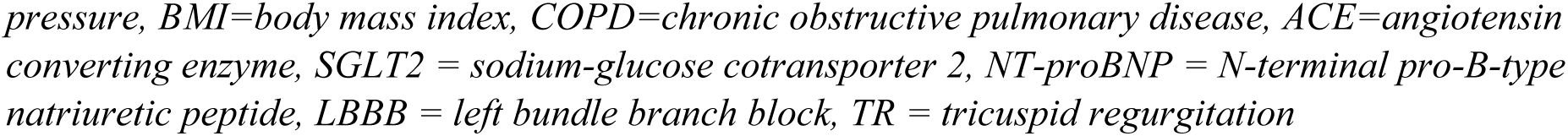
Baseline Clinical, Laboratory, Electrocardiographic, and Echocardiographic Characteristics Stratified By AI-ECG Diastolic Function Grade.

#### Clinical History and Comorbidities

HF history increased progressively across DF grades, present in approximately one-third of normal/Grade 1 patients and over half of Grade 3 patients. Atrial fibrillation was markedly more prevalent in Grade 3 (77.4%) than in normal/Grade 1 or Grade 2 groups (approximately 56%). Hypertension was common across all groups, with highest prevalence in Grade 2. Myocardial infarction and diabetes mellitus showed similar but less pronounced gradients.

#### Vital Signs and Laboratory Values

Systolic blood pressure was modestly higher in Grade 2, while BMI demonstrated a progressive decline across grades. Creatinine increased progressively, reflecting worse renal function in higher AI-ECG DF grades.

#### Medication Prescriptions

Baseline prescription of guideline-directed therapies exhibited consistent gradients. Beta-blocker prescription rose from approximately half of normal/Grade 1 patients to over two-thirds in Grade 3. ACE inhibitor/ARB use ranged from 40.0% to 49.0% across groups. Aldosterone antagonists use more than doubled from normal/Grade 1 to Grade 3. Use of ARNI and SGLT2-i, while low overall, was more frequent in grade 3.

#### Electrocardiographic Findings

Sinus rhythm predominated in normal/Grade 1 and Grade 2 groups (∼80%) but in only half of Grade 3 patients. QRS duration and LBBB prevalence increased progressively. Saliency mapping demonstrated that the AI-ECG model did not simply rely on these conventional ECG findings for DF grade determination (Supplemental Figure 3).

#### Echocardiographic Measures

Diastolic filling parameters, chamber dimensions, and systolic function differed markedly across groups. E/e′ ratio and indexed left atrial volume increased progressively with higher DF grades. LVEF demonstrated an inverse relationship, with median values declining from normal/Grade 1 to Grade 3; nearly half of Grade 3 patients had LVEF ≤40% compared with 15% in normal/Grade 1. Moderate-to-severe mitral and tricuspid regurgitation and elevated tricuspid regurgitation velocity were progressively more prevalent in higher DF grades.

### Disease Severity Markers

Among adjudicated cases with documented NYHA class, functional status varied across groups: NYHA III–IV was present in 32% of normal/Grade 1, 86% of Grade 2, and 94% of Grade 3 patients, although NYHA class was not recorded in much of the cohort. (Figure 2)

**Figure 2.**
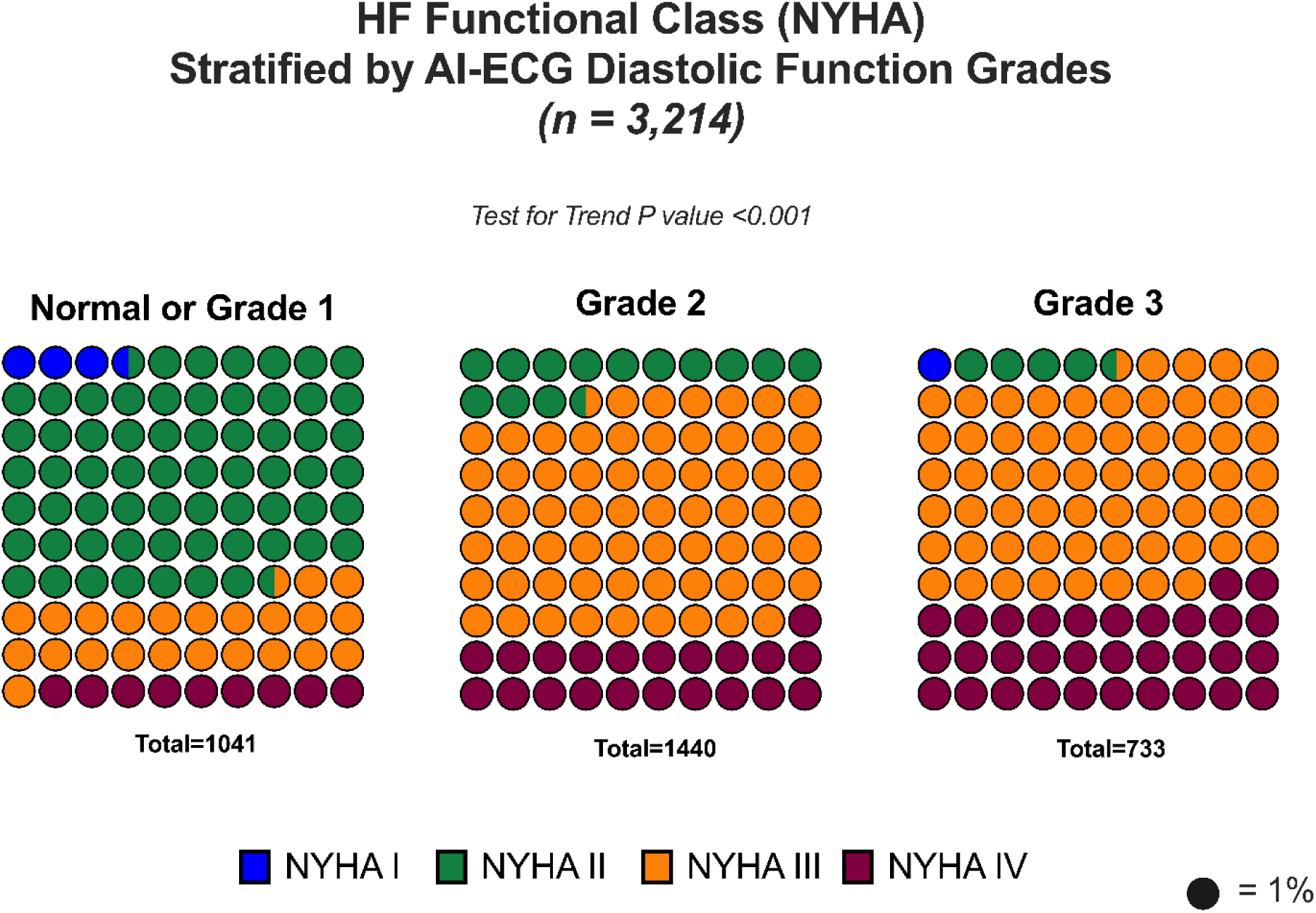
New York Heart Association functional (NYHA) Class Distribution Across AI-ECG Diastolic Function Grades Among Adjudicated Patients

NT-proBNP measurements were available in 8,201 patients (71.3%). Median NT-proBNP values increased across DF groups, from 2,172 pg/mL in the normal/Grade 1 group to 3,864 pg/mL in Grade 2 and 5,844 pg/mL in Grade 3 (P<0.001) (Figure 3A).

**Figure 3.**
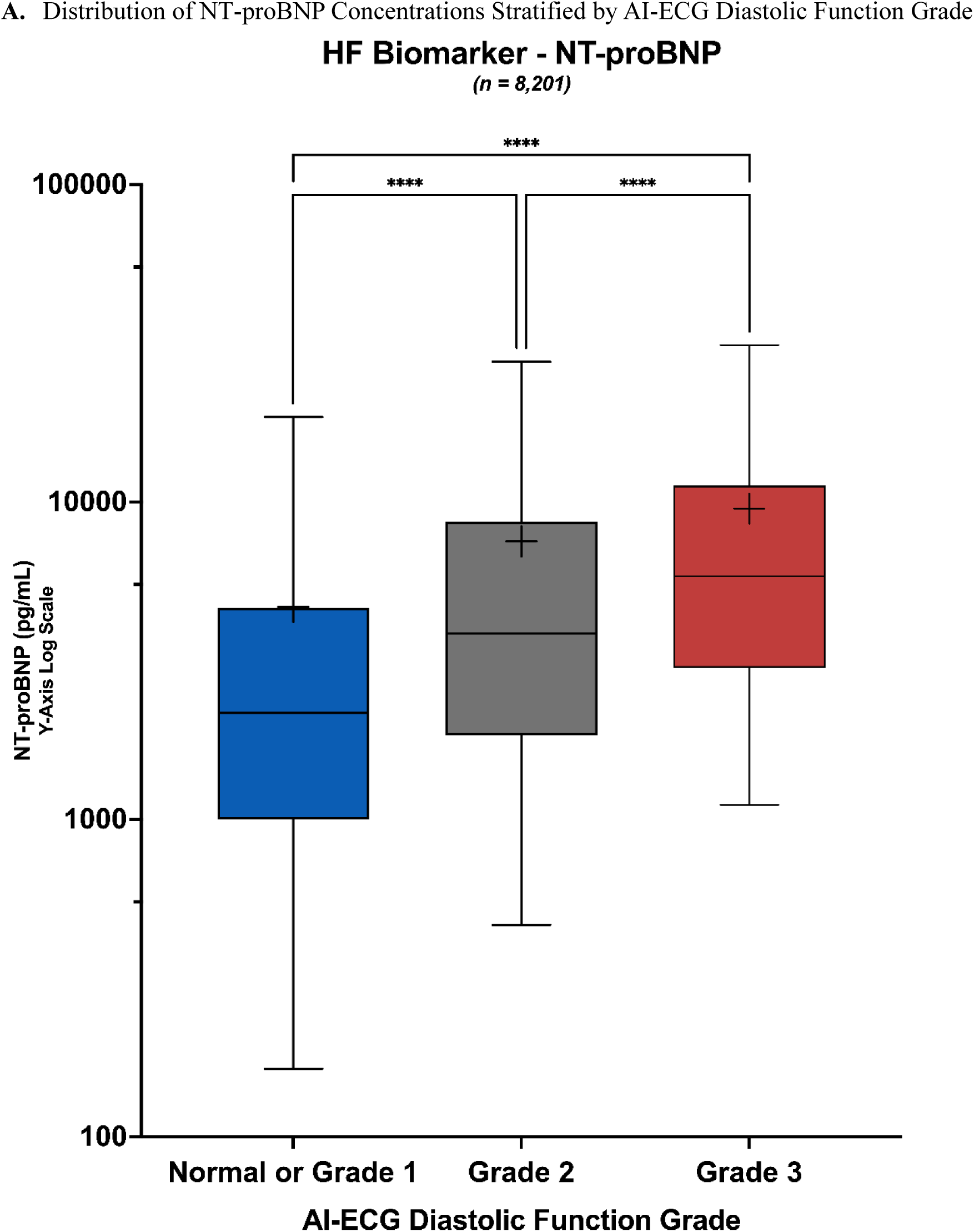
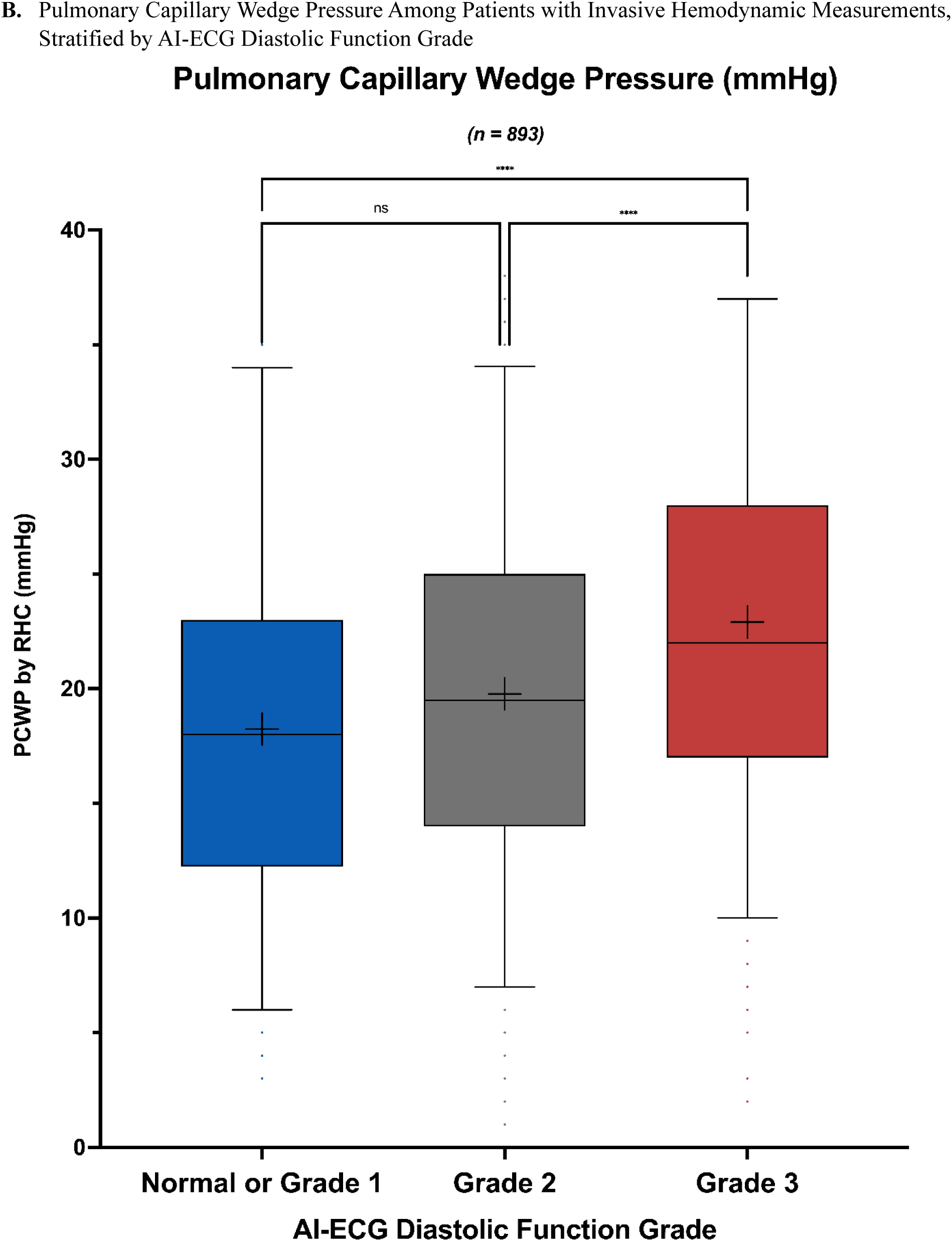
NT-proBNP Concentrations and Pulmonary Capillary Wedge Pressure by AI-ECG Diastolic Function Grade

Resting pulmonary capillary wedge pressure (PCWP) was available in 893 patients (7.8%), where baseline ECGs were typically within one day of the clinically indicated invasive study (overall range – 12.0 to +12.1 hours). Mean PCWP values were 18 (8.0) mmHg in the normal/Grade 1 DF group, 19.8 (8.2) mmHg in Grade 2, and 22.9 (8.3) mmHg in Grade 3. The comparison across the three groups was statistically significant (ANOVA P<0.001). In post-hoc analyses, Grade 3 differed significantly from both normal/Grade 1 (P<0.001) and Grade 2 (P<0.001), whereas the difference between normal/Grade 1 and Grade 2 did not reach statistical significance (P = 0.06). (Figure 3B)

Similarly, median MAGGIC risk scores incorporating NYHA class were higher in Grade 2 and Grade 3 compared with normal/Grade 1 (P<0.001).

### Clinical Outcomes

#### Overall Survival and Follow-up

Among the 11,513 patients in the analysis cohort, survival information was available for 11,511 patients, with 2 patients (0.02%) lacking survival status. 5,113 deaths occurred during the study period. The median survival time was 4.20 years, and the median follow-up among survivors was 3.68 years.

#### Index Hospitalization Length of Stay

The median length of stay for the index AHF hospitalization was 5.42 days (IQR, 3.63–8.49 days) in the overall cohort. Length of stay differed modestly across AI-ECG diastolic function categories (P<0.001), with median values of 5.7 days in the normal/Grade 1 group, 5.4 days in Grade 2, and 5.2 days in Grade 3.

#### Heart-Failure Rehospitalization

Across the cohort, 7,239 patients had a subsequent HF hospitalization recorded during follow-up. Among those who were re-hospitalized, the median time to HF rehospitalization differed significantly across DF categories (P<0.001). Median times were 119 days in the normal/Grade 1 group, 103 days in Grade 2, and 94 days in Grade 3. The cumulative incidence functions demonstrated progressive increases in HF rehospitalization and death across DF categories (Figure 4A).

**Figure 4.**
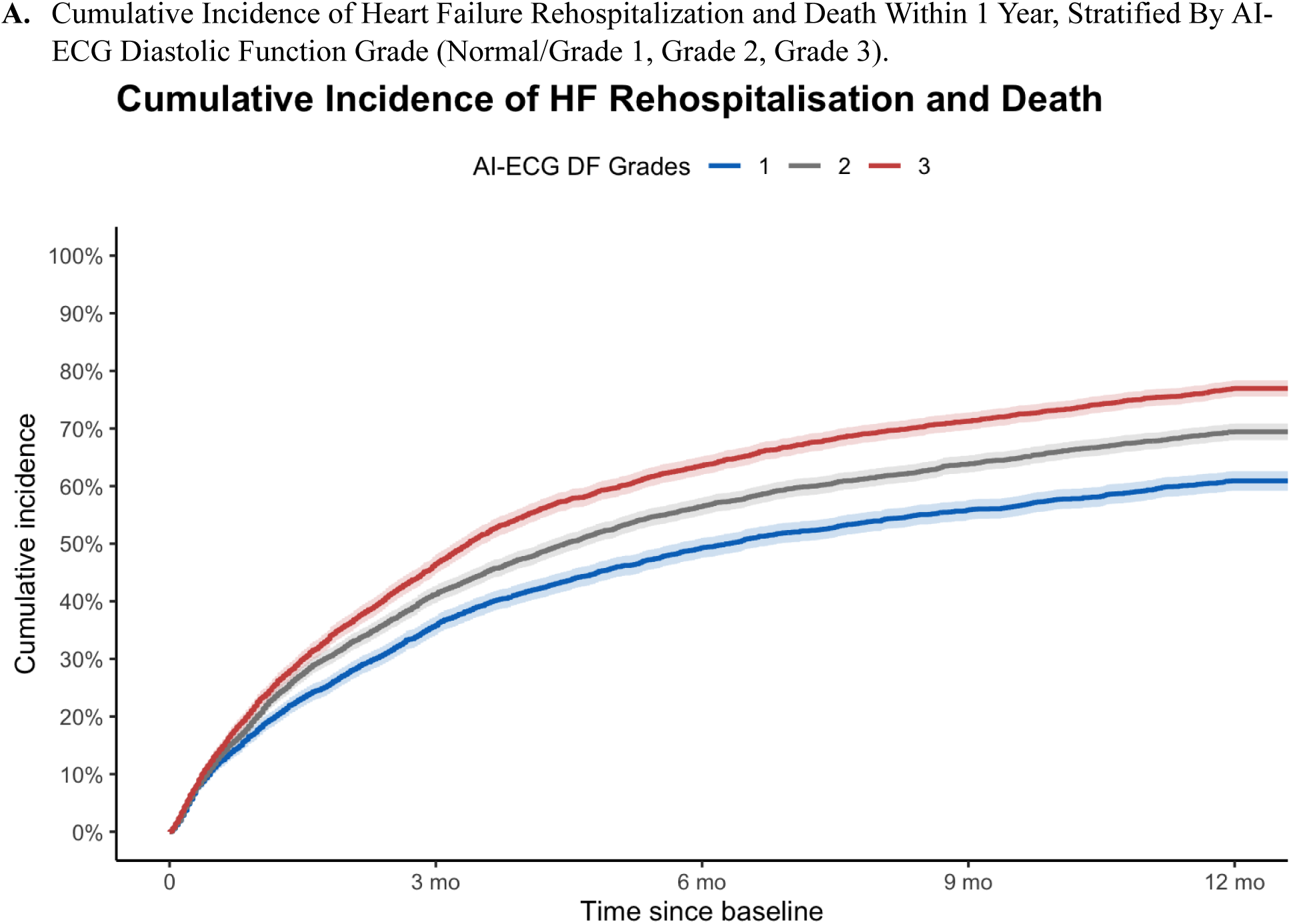
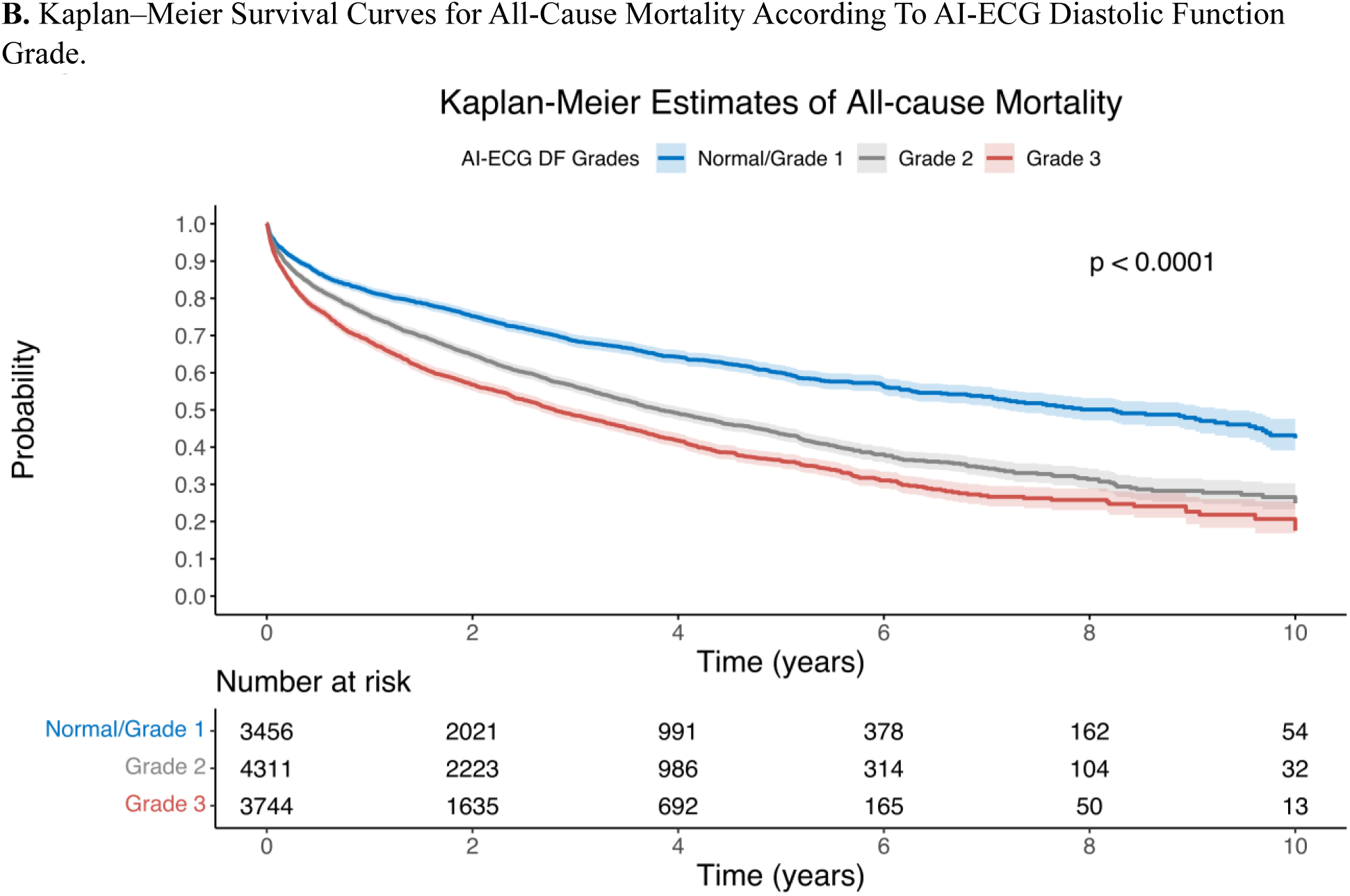
Heart Failure Rehospitalization and Survival by AI-ECG Diastolic Function Grade

### Survival Analysis

In the unadjusted Cox model, AI-ECG DF Grade showed a Graded association with mortality. Relative to the normal/Grade 1 group, HRs were 1.58 (95% CI, 1.47–1.70) for Grade 2 and 1.98 (95% CI, 1.84–2.13) for Grade 3 (C-index 0.57).

In adjusted multivariable Cox models (n = 11,383; 5,062 deaths), both AI-ECG–derived and echocardiography-derived DF grades were associated with mortality. Using the normal/Grade 1 AI-ECG DF group as the reference, the adjusted HR was 1.25 (95% CI, 1.16–1.35) for Grade 2 and 1.44 (95% CI, 1.33–1.56) for Grade 3 (C-index 0.71) (Supplementary Table 3). As shown in Figure 4B, survival differed significantly across AI-ECG DF grades (log-rank P<0.0001). Median survival declined progressively with worsening DF: 8.23 years (95% CI, 7.18–9.34) for the normal/Grade 1 group, 3.83 years (95% CI, 3.60– 4.15) for Grade 2, and 2.80 years (95% CI, 2.63–3.07) for Grade 3. In the echocardiographic DF model, compared with the normal DF reference group, the adjusted HR were 1.21 (95% CI, 0.88–1.67) for Grade 1, 1.45 (95% CI, 1.17–1.79) for Grade 2, 1.41 (95% CI, 1.11–1.78) for Grade 3, and 1.06 (95% CI, 0.85– 1.32) for indeterminate DF (C-index 0.70).

To evaluate whether combining traditional clinical risk with AI-ECG DF could enhance prognostic stratification, we created four groups defined jointly by the MAGGIC score (low vs high) and AI-ECG DF grade (normal/Grade 1 vs Grade 2–3). Patients with low MAGGIC scores (<22) and normal or Grade 1 DF (n = 2,593) served as the reference. Compared with this group, mortality was higher among those with low MAGGIC scores and Grade 2–3 DF (n = 4,257; HR, 1.6; 95% CI, 1.5–1.7; P<0.001). Risk increased further for patients with high MAGGIC scores (≥22) but normal or Grade 1 DF (n = 863; HR, 2.6; 95% CI, 2.3–2.9; P<0.001) and was greatest in those with high MAGGIC scores and Grade 2–3 DF (n = 3,799; HR, 3.4; 95% CI, 3.1–3.7; P<0.001). The proportional hazards assumption was met (P=0.17), and overall model discrimination was acceptable (C-index 0.62). Kaplan–Meier curves showed clear, ordered separation of the four strata, with poorest survival in the group with both high clinical risk and higher AI-ECG DF burden (Figure 5).

**Figure 5.**
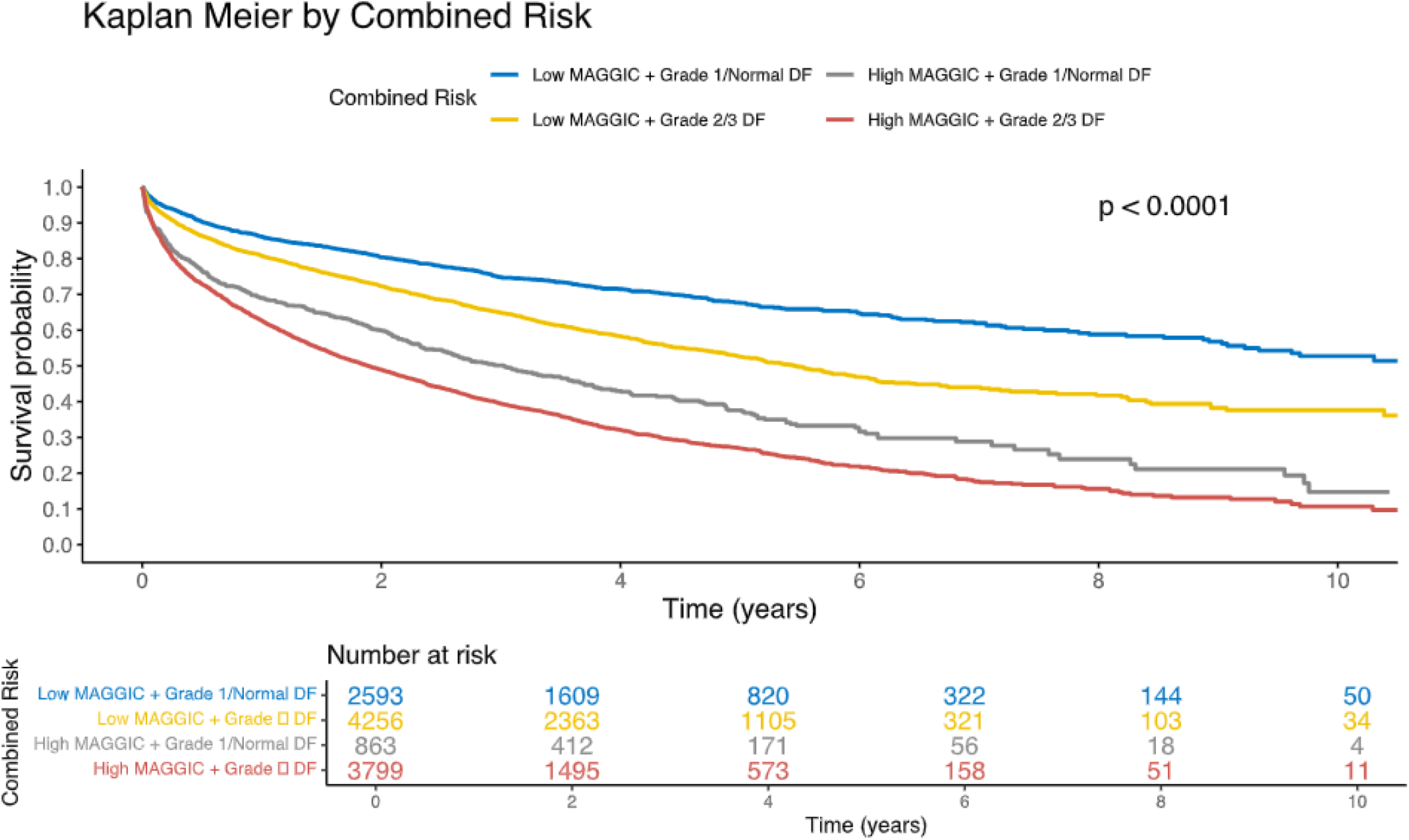
Kaplan–Meier Survival Curves for Combined Risk Stratification by MAGGIC Score and AI-ECG Diastolic Function Grade

### Sensitivity Analysis

NT-proBNP was excluded from the primary multivariable model due to substantial missingness in the cohort, but its established prognostic role warranted a sensitivity analysis in the subset with available measurements. In the NT-proBNP–adjusted model, AI-ECG DF grade was no longer independently associated with mortality. Compared with normal/Grade 1 DF, the HRs were 0.98 (95% CI, 0.90–1.07; *P* = 0.65) for Grade 2 and 1.04 (95% CI, 0.95–1.14; *P* = 0.37) for Grade 3. NT-proBNP demonstrated a strong association with mortality (HR 1.27 per log-unit; 95% CI, 1.23–1.31; P<0 .001), and associations for other covariates were consistent with the primary model.

In prespecified analyses, we evaluated whether the association between AI-ECG DF grade and mortality differed across clinically relevant subgroups. Interaction testing revealed no evidence of effect modification by reduced LVEF (≤40%; P-interaction = 0.27 for Grade 2 and 0.39 for Grade 3), sinus rhythm (P-interaction = 0.25 and 0.78), or indeterminate echocardiographic DF classification (P-interaction = 0.14 and 0.15), all exceeding the prespecified α of 0.01. Subgroup analyses were consistent with these results: as shown in the forest plot (Figure 6), higher AI-ECG DF grades were associated with increased mortality across all strata, with HR comparable to those observed in the overall cohort. These findings indicate that the prognostic association of AI-ECG DF grade was stable across LVEF, rhythm, and echocardiographic determinacy subgroups.

**Figure 6.**
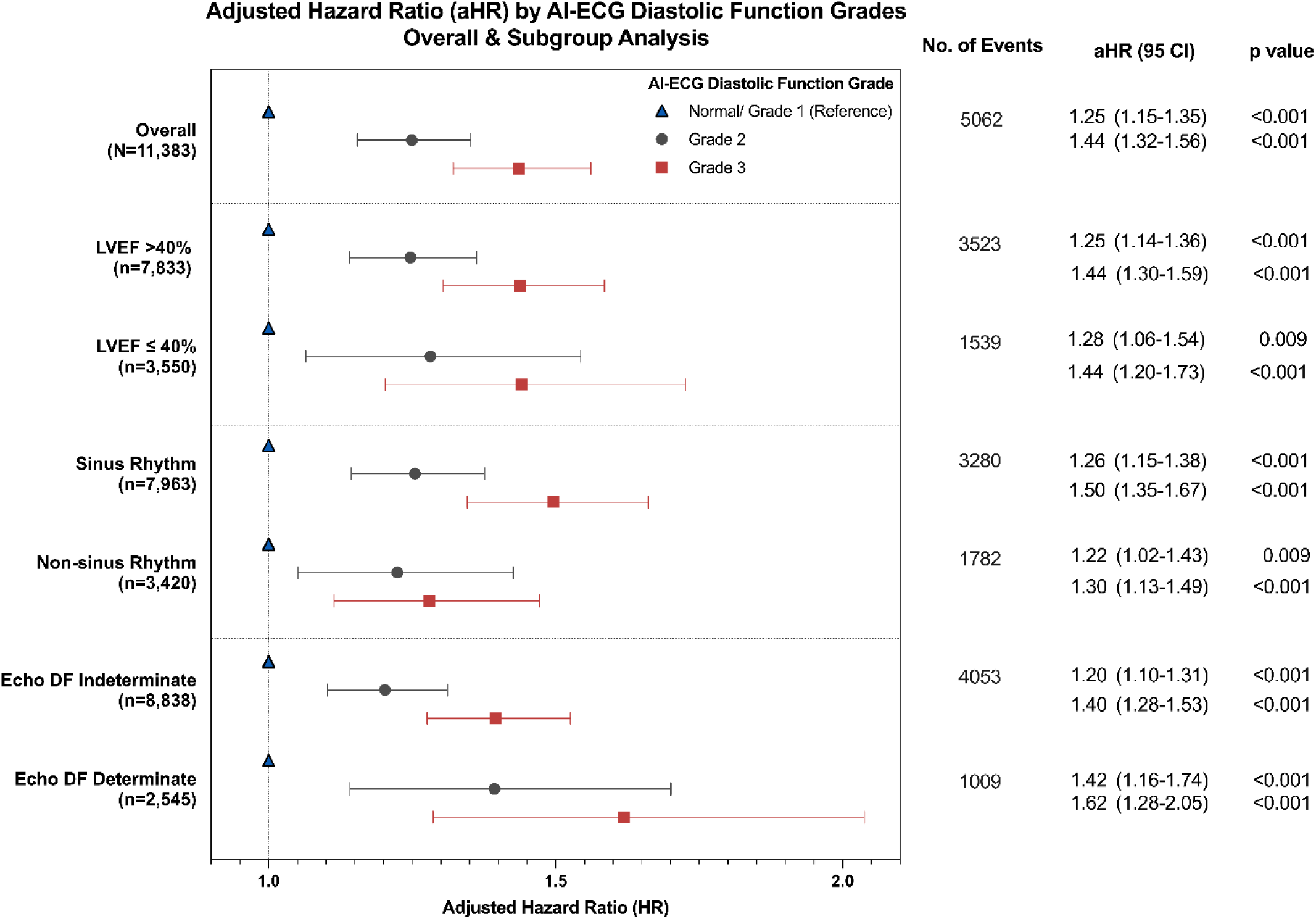
Adjusted Hazard Ratios of Main Survival and Subgroup Analysis for All-cause Mortality **Adjustment variables:** age, sex, body mass index (BMI), NYHA class, systolic blood pressure (SBP), sinus rhythm, QRS duration, left ventricular ejection fraction (LVEF), prior history of congestive heart failure (CHF), atrial fibrillation (AF), hypertension (HTN), myocardial infarction (MI), chronic pulmonary disease (COPD), diabetes mellitus (DM), current smoking, serum creatinine, sodium, hemoglobin, and baseline prescriptions of ACE inhibitor (ACEi), angiotensin receptor blocker (ARB), aldosterone antagonist (MRA), angiotensin receptor-neprilysin inhibitor (ARNI), and sodium-glucose cotransporter-2 inhibitor (SGLT2i).

### Longitudinal Change in AI-ECG–Estimated Filling Pressure Probability

Among 7,190 patients with serial ECGs (median 72 days post-discharge), estimated FP probability showed distinct temporal patterns across baseline AI-ECG DF grades. Repeated-measures ANOVA demonstrated a large effect of DF grade (F(2,7187)=6451.6; P<0.001) and a significant time × group interaction (F(2,7187)=639.5; P<0.001), whereas the overall mean change over time was small (F(1,7187)=8.89; P=0.003). Estimated marginal means illustrated these divergent trajectories. Normal/Grade 1 patients showed an increase in estimated FP probability from 0.54±0.01 to 0.69±0.01 (P<0.001), Grade 2 patients showed a decrease from 0.76±0.01 to 0.70±0.01 (P<0.001), and Grade 3 patients showed a slight decline from 0.86±0.01 to 0.83±0.01 (P<0.001). Between-group differences were significant at both baseline and follow-up (all P<0.001). (Figure 7A)

**Figure 7.**
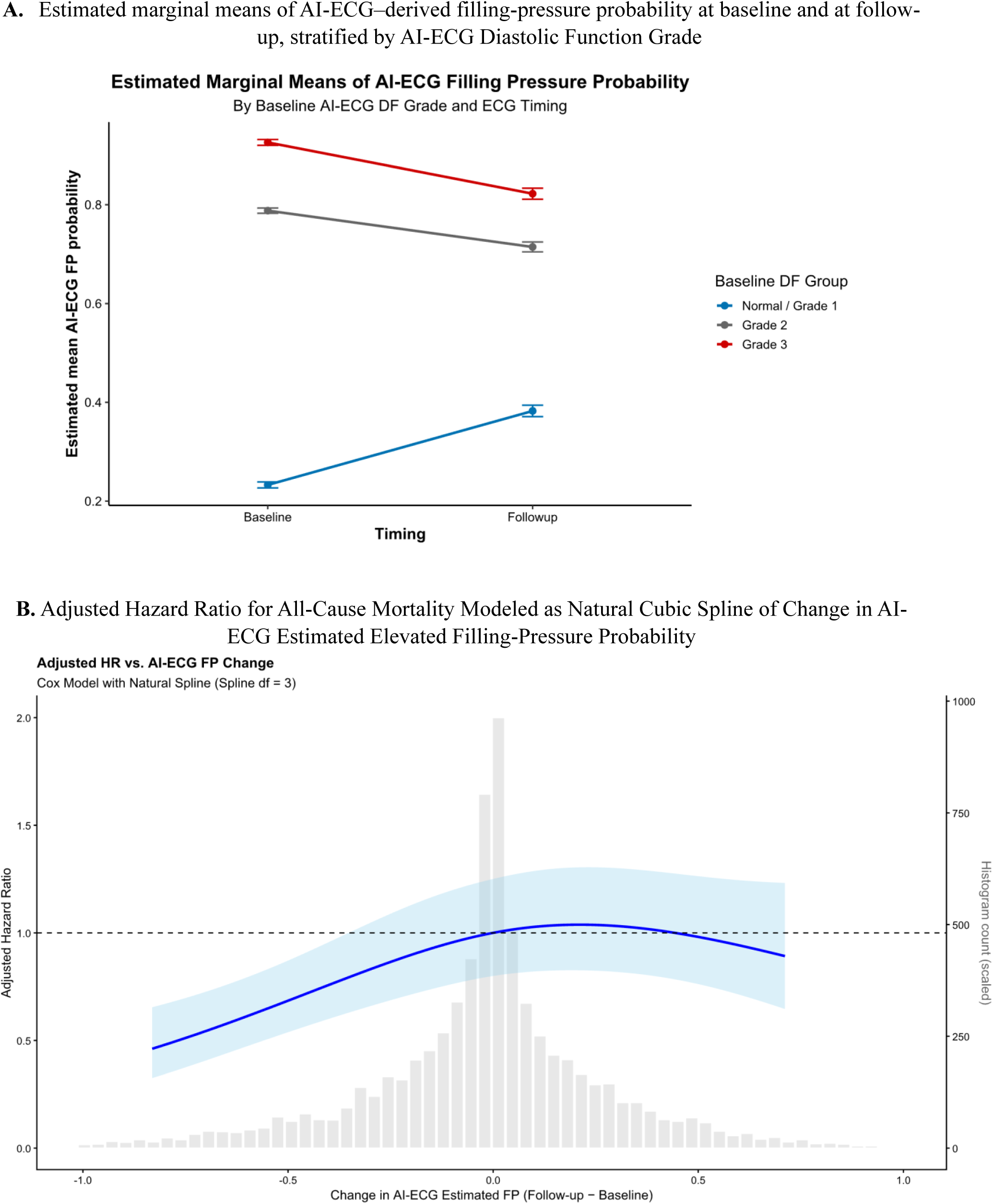
Longitudinal Change in AI-ECG–Estimated Elevated Filling-Pressure Probability and Association with Mortality

The distribution of change values centered around zero, prompting binary classification of improved versus unchanged/worsened patients. In a multivariable Cox model, improvement was independently associated with lower mortality (HR 0.85; 95% CI 0.79–0.91; P<0.001). To examine the full continuous relationship between FP-probability change and mortality, we modeled the change variable using a natural cubic spline. The spline terms and model fit indicated non-linear association (Δχ²=21.8; P<0.001). Improvement in FP probability corresponded to progressively lower adjusted risk, whereas the curve flattened around and beyond zero, revealing no statistically reliable increase in risk with worsening values (Figure 7B).

Overall, serial AI-ECG measures identified heterogeneous trajectories of estimated FP probability, and improvements were associated with reduced mortality, while worsening did not demonstrate a clear adverse gradient, beyond baseline AI-ECG DF grades. Importantly, these analyses were limited to a selected subset with repeat ECGs and should be interpreted as exploratory.

## DISCUSSION

In this large, geographically diverse, multisite AHF cohort, AI-ECG DF provided a practical, reproducible, noninvasive marker of hemodynamic severity. Higher DF grades were associated with older age, greater comorbidity burden, elevated natriuretic peptide, worse NYHA class, and higher MAGGIC risk scores. Echocardiographic and invasive hemodynamic markers showed progressive worsening across DF categories, accompanied by proportional differences in rehospitalization and mortality risk. AI-ECG DF remained independently associated with mortality after multivariable adjustment, consistent with prior observations in other disease states^14,15^. These findings indicate that AI-ECG DF reflects clinically meaningful variation in diastolic hemodynamic burden.

AI-ECG DF identified clinically distinct HF phenogroups among hospitalized AHF patients. Patients with normal or Grade 1 DF had lower natriuretic peptides, less structural remodeling, and the most favorable outcomes. Grade 2 DF was characterized by features consistent with HFpEF or HFmrEF, including hypertension, left atrial enlargement, elevated E/e′ ratio, and intermediate MAGGIC risk. Grade 3 DF identified patients with advanced structural disease, including marked atrial and ventricular remodeling, more significant valvular regurgitation, higher tricuspid regurgitation velocity, higher AF prevalence, and reduced LVEF in almost half of the cohort. Furthermore, index hospitalization length of stay did not differ across DF grades despite markedly different hemodynamic burden and outcomes, suggesting advanced diastolic dysfunction is not easily recognized during routine care. Clinicians may discharge patients once symptoms and intravascular hypervolemia improve, even when filling pressures remain elevated. AI-ECG DF may identify this hidden risk. These findings align with population-based data demonstrating that AI-ECG DF materially improves HF risk discrimination when added to established risk models. Desai et al. showed that adding AI-ECG DF to PREVENTHF risk score materially improved near term HF risk discrimination and reclassified risk, with a 10–20 fold higher incident HF risk among screen-positive individuals.^16^ Consistent with this, nearly half of our AI-ECG Grade 3 patients had no prior HF diagnosis documented, underscoring the diagnostic value of AI-ECG DF in incident HF presentations. Collectively, these observations support the potential utility of AI-ECG DF across the LVEF and HF stage (B to D) spectrum.

In our cohort, AI-ECG DF confers a clear prognostic signal. Relative to patients with normal or Grade 1 AI-ECG DF, Grade 2 was associated with 25% higher adjusted mortality risk and Grade 3 with 44% higher risk. Beyond clinical risk, AI-ECG DF provided incremental risk stratification in addition to established risk scores, such as the MAGGIC score. Among patients with low MAGGIC score, Grade 2–3 DF is associated with 60% higher risk of death compared to normal/Grade 1 DF, and 240% higher when both high MAGGIC score and Grade 2–3 DF were present, with clear divergent of survival risks. In sensitivity analyses limited to those with natriuretic peptides, adjustment for NTproBNP attenuated the independent association of AI-ECG DF with mortality, while NTproBNP remained prognostic. We hypothesize that this attenuation is biologically coherent as both potentially measure myocardial stretch along the same pathophysiologic pathway but from different vantage points: NTproBNP as a soluble peptide marker of wall stress and AI-ECG DF as an electromechanical signature of elevated FP. Thus, NT-proBNP likely mediates part of the risk conveyed by AI-ECG DF, supporting complementary clinical utility of these biomarkers.

The risk profile of AI-ECG Grade 3 patients, with median survival of 2.8 years, was comparable to high-risk HF trials and registry populations. VICTORIA-HF, which evaluated the efficacy of vericiguat, is widely recognized as one of the sickest chronic HFrEF trial populations in recent years. In that study, projected median survival was 2.5 to 3 years.^17^ COAPT, evaluating transcatheter mitral valve repair in secondary MR, reported 46% 2-year mortality in the control arm^18^. Similarly, CHAMPION enrolled NYHA class III patients with a recent HF hospitalization and used implantable pulmonary artery pressure monitoring to guide therapy; the trial documented comparable high event rates in the control arm and showed that hemodynamic-guided care reduced recurrent decompensation^19^. In the HELP-HF registry, patients meeting all HFA–ESC advanced HF criteria experienced 47% one-year mortality.^20^ These comparisons suggest AI-ECG Grade 3 identifies patients with risk trajectories warranting HF management intensification or specialist referral. Determining whether AI-ECG DF can facilitate earlier recognition of such individuals will require prospective evaluation.

The longitudinal analyses provided additional insight into the hemodynamic trajectories captured by AI-ECG FP probability after discharge. Patients with Grade 2 and Grade 3 DF demonstrated modest reductions in estimated FP during follow-up, a pattern consistent with overt congestion being recognized during hospitalization and aggressively treated. In contrast, patients with normal or Grade 1 DF showed gradual increases in filling-pressure probability, suggesting the presence of milder or predominantly interstitial congestion that may be less apparent clinically and therefore less intensively addressed. This divergence mirrors established physiologic differences between HF phenotypes: HFpEF tends to exhibit greater interstitial fluid accumulation with relatively preserved intravascular volume, whereas HFrEF more often demonstrates substantial intravascular expansion; in both phenotypes there is incomplete normalization of congestion at the time of discharge despite clinically effective diuresis.^21^ The latter further exemplifies the importance of post-discharge DF assessment and monitoring. The independent association between improved FP probability and outcomes favors a biological phenomenon beyond regression to the mean.

Recent commentary emphasizes that most AI tools fail to deliver clinically meaningful signals, add workflow burden, or lack real-world utility evidence.^22^ These concerns fuel ongoing skepticism around the expanding catalog of AI-enabled biomarkers. In contrast, this analysis demonstrates characteristics of a potentially high-value AI biomarker. First, AI-ECG DF captures a physiologically plausible hemodynamic signature: associations with clinical severity measures suggest the model identifies biologically relevant myocardial stretch patterns rather than spurious correlations. Second, prognostic gradients extend across short to long-term horizons and remain consistent across rhythm, EF, and echocardiographic determinacy subgroups, indicating broad applicability. Third, using routine 12-lead ECG during hospitalizations, the approach requires no new hardware and integrates seamlessly into existing workflows. Finally, the ability of serial FP probability trajectories to identify heterogeneity in post-discharge risk aligns with emerging emphasis on longitudinal digital phenotyping. These features suggest that AI-ECG DF functions less as “yet another algorithm” and more as a biologically plausible, deployable diastolic biomarker that may meaningfully complement clinical assessment in AHF.

### Limitations

Several limitations merit consideration. First, this retrospective cohort from a single health system, though spanning multiple geographic regions and practice settings, requires external validation in independent AHF cohorts. Second, requiring both ECG and TTE during hospitalization enriches the cohort and potentially limits generalizability. Third, echocardiographic DF was unavailable or indeterminate in a substantial fraction, reflecting real-world constraints but limiting direct comparison between AI-ECG and echo DF across the full cohort. Fourth, residual confounding remains possible despite extensive multivariable adjustment, as important prognostic determinants including frailty, socioeconomic status, adherence, and detailed device history were incompletely captured. Fifth, natriuretic peptides were unavailable in a sizable proportion, influencing our modeling strategy. Furthermore, non-cardiac conditions raising NT-proBNP (sepsis, renal failure, pulmonary embolism) confound observations. Hence, attenuation of AI-ECG DF’s independent mortality association in NT-proBNP-adjusted analyses should be interpreted cautiously. Finally, serial ECGs during follow-up were obtained through routine care rather than structured protocol and may reflect illness severity, introducing informative censoring. Hence, longitudinal analyses are hypothesis-generating and require prospective confirmation with standardized follow-up.

## CONCLUSION

Across a large, multisite AHF population, AI-ECG–derived diastolic function Grades aligned with biomarker, echocardiographic, and invasive measures of disease severity and were associated with adverse outcomes in the short, intermediate, and long term. Distinct DF phenogroups, defined by physiological and prognostic associations observed, showed unique post-discharge trajectories that clarify congestion states and recovery after AHF hospitalization. Overall, AI-ECG DF may function as a complementary biomarker to refine hemodynamic assessment and identify patients requiring intensified monitoring or specialist referral.

## Data Availability

The data that support the findings of this study are not publicly available due to information that could compromise the privacy of research participants. Requests for data sharing will be considered on a case-by-case basis and are subject to Mayo Clinic data sharing policies and institutional review board approval.

